# XGBoost, a novel explainable AI technique, in the prediction of myocardial infarction, a UK Biobank cohort study

**DOI:** 10.1101/2022.04.08.22273600

**Authors:** Alexander Moore, Max Bell

## Abstract

**Background and objective:** Myocardial infarction is common and associated with high morbidity and mortality. This study assessed if “Explainable AI” in the form of extreme gradient boosting (XGBoost) could outperform traditional logistic regression in predicting myocardial infarction (MI) in a large cohort.

**Methods:** We compared two machine learning methods, XGBoost and logistic regression in predicting risk of MI. The UK Biobank is a population-based prospective cohort including 502 506 volunteers with active consent, aged 40–69 years at recruitment from 2006 to 2010. These subjects were followed until end of 2019 and the primary outcome was myocardial infarction.

**Results:** Both models were trained using 90% of the cohort. The remaining 10% was used as a test set. Both models were equally precise, but the regression model classified more of the healthy class correctly. XGBoost was more accurate in identifying individuals who later suffered a myocardial infarction. Receiver operator characteristic (ROC) scores are class size invariant. In this metric XGBoost outperformed the logistic regression model, with ROC scores of 0.86 (accuracy 0.75; precision 0.99 and recall 0.75 for the healthy class; precision 0.07 and recall 0.81 for the MI-class) compared to 0.77 (accuracy 0.77; precision 0.99 and recall 0.77 for the healthy class; precision 0.07 and recall 0.78). Secondly, we demonstrate how SHAPley values can be used to visualize and interpret the predictions made by XGBoost models, both for the cohort test set and for individuals.

**Conclusions:** The XGBoost machine learning model shows very promising results in evaluating risk of MI in a large and diverse population. This model can be used, and visualized, both for individual assessments and in larger cohorts. The predictions made by the XGBoost models, points towards a future where “Explainable AI” may help to bridge the gap between medicine and data science.

## Introduction

In adults, the prevalence of cardiovascular disease (CVD) is close to 50% and is a major cause of morbidity and mortality worldwide.[1] Notably, this holds true even though annual death rates attributable to coronary heart disease (CHD) declined 31.8% from 2006-2016.[1] Similarly, a study of over 45 000 hospitalizations for myocardial infarction (MI) showed a 24% decrease from 2000-2008.[2]

This substantial decline in the incidence of coronary heart disease is partly driven by modifiable risk factors; changes in cholesterol, improved blood pressure control, decreased smoking and increased physical activity.[3] Correctly focusing primary preventive efforts, such as lifestyle changes, and identifying the patients who stand to benefit the most from preventative medication requires an understanding of the risk factors for the development of CVD and MI. This was clearly shown in the ASPREE study, where over 19,000 elderly patients were randomly assigned to oral aspirin or placebo.[4] No decrease in CVD was seen in the aspirin group, on the contrary more severe bleeding events were observed, highlighting the benefits of individualized risk stratification.

Multiple risk factor models exist. Following the Framingham Risk Score, the first large study of CVD risk factors,[5] risk assessment tools from the American and European cardiology societies have been launched; the EURO Score[6] and the American College of Cardiology (ACC)/American Heart Association (AHA) Heart Risk Score.[7] These scores include classic risk factors such as age, gender, diabetes, smoking and blood pressure. To increase accuracy and individualization for cardiovascular risk prediction, biomarkers, such as blood lipids have been added to the Framingham Risk Score and to the EURO Score.[6] Recently, high-sensitivity cardiac troponins have been shown not only to be able to detect acute MI, but to function as predictors of adverse outcomes.[8, 9, 10] The Norwegian HUNT study added high-sensitivity C-reactive protein, troponins and cholesterol to above mentioned classic risk factors.[11] Specifically, troponins provided improved prognostic information.

With or without adding biomarkers, an alternative path to the improvement of predictive properties is opened by the development of machine learning (ML). Recently, a machine learning algorithm predicted the likelihood of acute MI, incorporating age, sex, with paired high-sensitivity cardiac troponin I outperformed the European Society of Cardiology 0/3-hour pathway.[12] This ML algorithm was trained on 3013 patients and tested on 7998 patients with suspected myocardial infarction.[12]

The present study aims to test a machine learning model, XGBoost[13] on predicting myocardial infarction in a population-based cohort of over 500 000 subjects, the UK Biobank.[14] We hypothesized that the ML algorithm would outperform logistic regression models.

## Methods

### Study design

This was a prospective observational study, testing machine learning and traditional logistic regression models, described in detail below. The North West Multi-Centre Research Ethics Committee approved the UK Biobank study. All participants provided written informed consent to participate. This research has been conducted using the UK Biobank Resource under Application Number 54045.

### Data source, the UK Biobank

UK (United Kingdom) Biobank is a large, population-based prospective study, established to allow detailed investigations of the genetic and non-genetic determinants of the diseases of middle and old age.[15, 16] The 500,000 participants were assessed from 2006 to 2010 in 22 assessment centres throughout the UK, the range of settings provided socio-economic and ethnic heterogeneity and an urban–rural mix. The wide distribution of all exposures allows generalizable associations between baseline characteristics and health outcomes. The assessments consisted of electronic signed consent; a self-completed touch-screen questionnaire; a computer-assisted interview; physical and functional measures and the collection of blood, urine, and saliva.

### Patient and Public Involvement statement

The investigation was conducted using the UK Biobank resource. Details of patient and public involvement in the UK Biobank are available online. No patients were involved in the research question or the outcomes, nor were they involved in design the study. No patients advised or interpreted results. There are no specific plans to disseminate the results of the present project to UK Biobank participants, but the UK Biobank disseminates key findings from projects on its website.

### Primary Outcome

Our models were trained to predict myocardial infarction. This was extracted from data-field 6150 in the UK Biobank (“vascular/heart problems diagnosed by a doctor”). There have been three separate instances where this question was posed to the participants (initially between 2006-2010, the first follow up was between 2012-2013 and the final follow up was in 2014). Any participant who selected “Heart Attack” (11849 participants) when answering this question in any of these instances was included in our positive class. More details on how these features are defined can be found through the UK Biobank.[15]

### Logistic Regression

Logistic regression models are commonly used for risk factor analysis.[17] They use a logistic function to model a binary dependent variable. A linear combination of predictors (input features) is used to calculate the probability that a given input belongs to one of the two classes. As features are combined linearly, explaining the predictions made by logistic regression models is straightforward. Moreover, logistic regression models have been shown to produce classification accuracies that are comparable to state of the art machine learning techniques.[18] In this study we implement a linear model for regularized logistic regression.

### XGBoost

Extreme Gradient Boost (XGBoost) is a powerful ensemble learning method, well suited to tabular datasets. Ensemble learning methods aggregate predictions of many individually trained classifiers, the combined prediction is typically more powerful than an individual classifier.[19] In the case of XGBoost the ensemble’s constituent classifiers are decision trees. Decision trees are graphical models, where distinct nodes are connected by branches, they are powerful tools for classification.[20] Each node represents a condition that is used to split the data. Data will pass along different branches dependent on whether it satisfies the condition at a particular node. Nodes at the bottom of a decision tree are known as leaf nodes. During training a tree adjusts the decision rules of its nodes in order to maximally separate the training data[21]. The most common class of the training data that terminate in a leaf node is assigned to that node after training. In order to classify new data, it must first be passed down the decision tree. Its class is then determined by the class assigned to the leaf node it terminates in.

XGBoost combines decision trees using a process known as gradient boosting.[13] Boosting methods build classifiers sequentially, such that the error from one classifier is passed on to the next. By training decision trees on the gradient of the loss produced by the previous tree, XGBoost is capable of producing prediction accuracies that match many state of the art supervised learning techniques, including neural networks.[22] In machine learning hyperparameters configure various aspects of an algorithm, they must be set before training and they can have a big impact on performance.[23] There are several hyperparameters controlling XGBoost, for example the number of decision trees to include must be specified. To maximise the accuracy of XGBoost these hyperparameters must be optimized. In order to maximise the utility of the training set, hyperparameter optimization typically uses a process known as cross-validation. Cross-validation randomly partitions the training set to produce a small validation set, used to measure performance. In this way, the effect of changing a hyperparameter can be quantified and the optimal value can be selected. Multiple rounds of cross-validation, with different partitions, are common.[20]

The following hyperparameters are typically optimised before training an XGBoost model: the number of estimators (decision trees), the max depth of a given decision tree, the minimum child weight in a decision tree, the minimum loss reduction required to make a partition (gamma), the number of columns to be subsampled when constructing a tree, and a regularization parameter (alpha).

Shapely values (SHAP values) originated as a concept in 1951 from cooperative game theory.[24] More recently they have been used as a tool for interpreting ensemble tree models. They facilitate the explanation of highly non-linear models, such as XGBoost, breaking down the impact of input features on prediction.[25] The SHAP value of a feature is calculated using the change in a model’s output if that feature’s value was replaced with a baseline value.[26] Consequently, considering the sum off all SHAP values is equivalent to considering the overall difference between a model’s prediction and the baseline.[26] In addition to breaking down the importance of an individual’s input features, SHAP values can explain the global impact of features across a population. SHAP values can be visualised in a number of ways.[26]

### Model training and validation

When validating supervised learning algorithms, it is essential that a portion of the data (∼10%) is set aside and not used in training. This data is known as the test set. An algorithm’s performance can be assessed by measuring its ability to correctly map inputs to outputs in the test set. Measuring performance on data used during training can result in overfitting or selection biases.[27]

Crude accuracy metrics, the total number of correct classifications, can be misleading when assessing the performance of a classification model. Consider a classification task where 0.01% of the population are high risk, a model could achieve 99.99% accuracy by classifying all individuals as low risk. Receiver operator characteristic (ROC) scores reflect the area underneath a curve obtained by plotting the true positive rate against the false positive rate. They are a more useful measure of performance in classification tasks with unbalanced classes.

## Results

### Model Performance

Results comparing XGBoost with a logistic regression model are presented in Table 1. The models were trained using 90% of the cohort (∼450,000 participants). The remaining 10% (∼50,000 participants) was set aside for use as a test set. The scores in Table 1 reflect the performance of both models when classifying the test set.

**Table 1.**

|  | XGBoost |  | Logistic Regression |  |
| --- | --- | --- | --- | --- |
|  | ROC Score 0.86 |  | ROC Score 0.77 |  |
|  | Accuracy 0.75 |  | Accuracy 0.77 |  |
| Class: | Precision | Recall | Precision | Recall |
| 0.0 (Healthy) | 0.99 | 0.75 | 0.99 | 0.77 |
| 1.0 (MI) | 0.07 | 0.81 | 0.07 | 0.78 |

Both models are equally precise but deviate when it comes to recall. The logistic regression model has a slightly higher recall for class 0.0 (the healthy class) whereas XGBoost has a higher recall for class 1.0 (the MI class). This means that the logistic regression model classified more of the healthy class correctly, whilst XGBoost was more accurate when it came to identifying individuals who would go on to suffer a myocardial infarction (MI). The above explains why the logistic regression model has a slightly higher crude accuracy score, as the healthy is significantly larger than MI class. A model that is more accurately recalling participants in the larger class is likely to end up with a larger total number of correct classifications.

ROC scores are class size invariant. In this metric XGBoost scores significantly higher than the logistic regression model (+0.09).

### Model Visualisation

SHAP values from an XGBoost model trained to predict MI using the UK Biobank cohort are shown in Figure 1. These were calculated for the 50,000 individuals in the test set. Each of these individuals is represented by a single data point. Here we see that having a high feature value for sex (being male) has a positive SHAP value, which means that being male increased the likelihood of being classified as class 1.0 (MI class). Conversely, a low feature value for sex (being female) has a negative impact on the model, meaning being female contributed to being classified as class 0.0 (healthy class). Features in Figure 1 are ordered based on their cumulative effect on model output.

**Figure 1.**
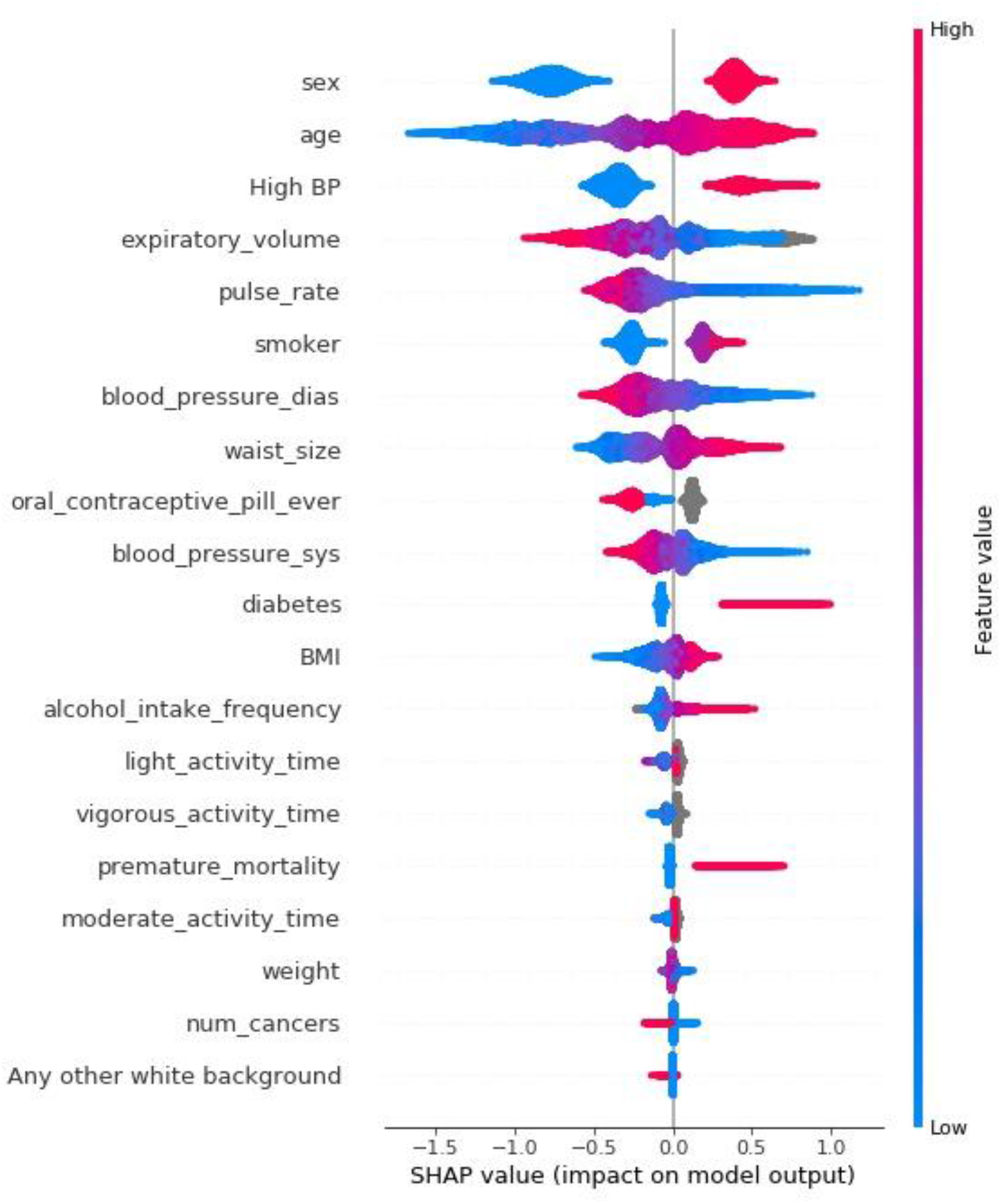
SHAP values extracted from a XGBoost model trained to predict MI (∼50,000 participants)

SHAP values can also be used to unpack individual predictions made by XGBoost. In Figure 2, the SHAP values for two individuals are displayed, one who was classified as MI class (2i) the other was classified as healthy (2ii). Large positive SHAP values for diastolic blood pressure and pulse rate, seen in Figure 2i, contributed to one individual being classified as MI class. These features have large negative SHAP values for the other individual, seen in Figure 2ii, meaning they strongly contributed to the individual being classified as healthy. Once again, features are ordered based on their impact on model output. The ordering of features is different in Figure 2i and 2ii as the relative importance of input features can vary between individuals.

**Figure 2.**
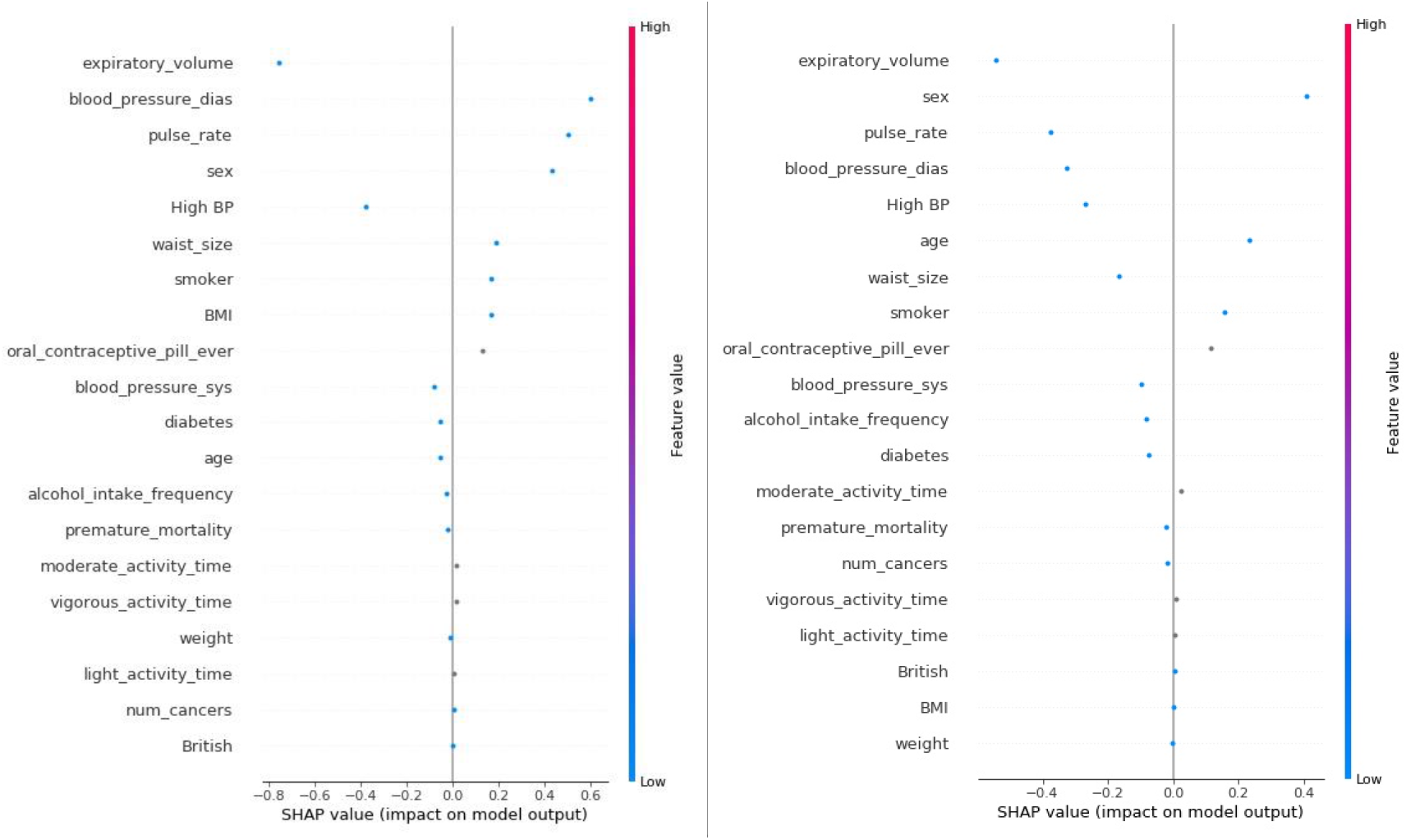
Figure 2i: SHAP values for an individual classified as MI class Figure 2ii: SHAP values for an individual classified as healthy class

A SHAP dependence plot represents the impact of two input variables on classification. Figure 3 explores the impact of waist size and sex on MI. Each data point represents an individual in the test set, colour coded by their sex. Waist size is plotted on the x-axis and the SHAP values (or model impact) are plotted on the y-axis. From inspecting Figure 3 we can see that most of the individuals with particularly small waists are female, this is associated with negative SHAP values. In these cases, an individual’s waist size made it more likely that they would be classified as healthy. As waist size increases SHAP values increase, this relationship continues linearly to a large cluster of individuals with positive SHAP values. In this cluster waist size made it more likely that an individual would be classified as belonging to the MI class. The positive correlation between waist size and SHAP value does not continue beyond the large cluster.

**Figure 3:**
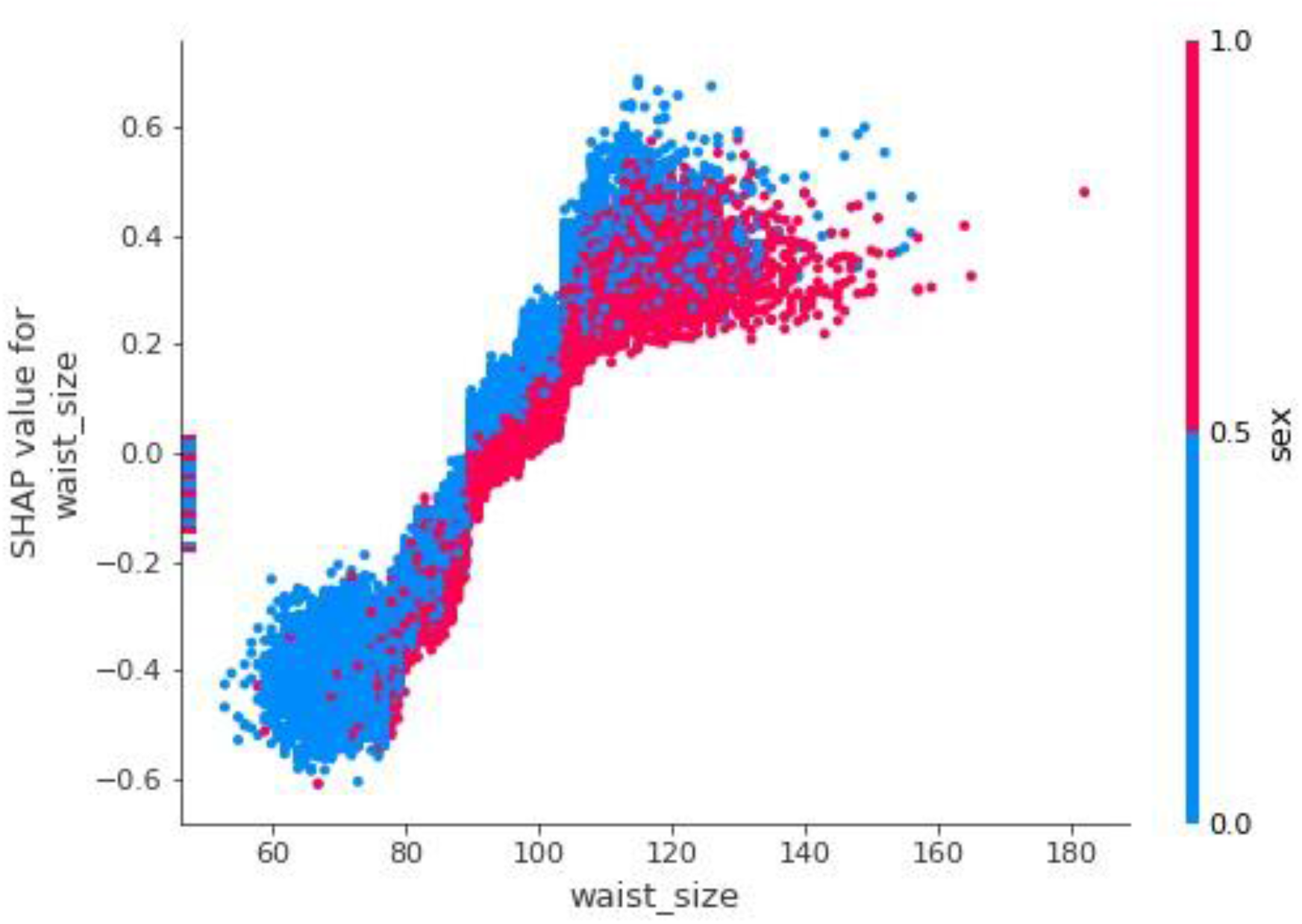
A SHAP dependence exploring the relationship between waist size, sex and myocardial infarction

The impact of having an extremely large waist is different for men and women. The men with the largest waists have SHAP values that are comparable with individuals in the cluster, implying their waist size had no additional impact on classification. On the other hand, women in the large cluster tend to have the highest SHAP values; waist size is likely to play a bigger role in classification for these women.

## Discussion

In this large cohort, the XGBoost machine learning model outperformed a multivariable logistic regression model in assessing risk of myocardial infarction. Importantly, our XGBoost model also allows for personalized risk estimations.

Previous attempts at using ML models exist in the field of cardiology. In 2005, Green and co-workers trained artificial neural network (ANN) ensembles and logistic regression models on data from 634 patients presenting in the emergency department with chest pain.[28] The ANN model performed slightly better than the logistic regression, but this was a single center study with limited power. Another small study, using data from 310 patients, tested ANN-algorithms for early diagnosis of acute myocardial infarction and prediction of size of infarction in patients presenting with chest pain.[29] The authors conclude that specifically designed ANN-algorithms allow early prediction of major AMI size and could be used for rapid assessment of these patients. Numerous studies outside of cardiology have tested machine learning models with varying degrees of success. Deep learning systems had high sensitivity and specificity for identifying diabetic retinopathy and related eye diseases.[30] Using machine learning technology to correctly classify indeterminate pulmonary nodules increased the reclassification performance, as compared to that of existing risk models.[31] In contrast, ML algorithms did not outperform traditional regression approaches in a low-dimensional setting for outcome prediction after traumatic brain injury.[32]

The findings in the present study have multiple implications. First, we show that ML-algorithms allow prediction of MI-risk in a large population. In future studies, when more high-resolution data is added, it is likely that the machine learning models will outperform logistic regression by a wider margin. Secondly, the XGBoost model predicts individual risk: this allows for patients with elevated risk patterns to aim for targeted and tailor-made life-style changes and in some cases medical treatments. In concert, these personalized interventions could decrease cardiac morbidity and possibly even add life years. As seen in the results section; the logistic regression model had a slightly higher crude accuracy score, as the healthy participants were significantly more common than individuals who would go on to suffer a myocardial infarction. Any model that is more accurately recalling participants in the larger class is likely to end up with a total number of correct classifications. In contrast, ROC scores are not affected by variable class sizes. In this metric XGBoost scored significantly higher than the logistic regression model. This means the risk of false negatives (a person at high risk of MI is classified as safe) is minimized using XGBoost; from an individual and epidemiological standpoint the cost of false negatives is much higher than false positives.

Our study has strengths and limitations. We had access to a large high-resolution longitudinal dataset from the UK Biobank. We trained the XGBoost ML model on 450 000 subjects and tested its performance on 50 000 individuals; compared to previous machine learning cardiology studies with less than 1000 patients, this adds to both model performance and generalizability. Comparing traditional logistic regression with ML models adds to transparency regarding the utility of these novel techniques. Said transparency is further achieved by choosing XGBoost – this is not a “black box” ML system; we can follow how the data flows through the system. Moreover, we demonstrate how XGBoost lowers misclassification events of patients at risk of myocardial infarction. A potential limitation to any MI-study is how the event is classified and recorded. If we had full biomarker and ECG data on all UK Biobank participants, we would likely have many more cases; that would enable more accurate machine learning and logistic regression predictions. Moreover, we acknowledge that our logistic regression model could be improved by adding interaction terms and accounting for non-linearity with regards to certain inputs. It is certainly possible to create more precise logistic models, but this requires active input from both statisticians and physicians with experience in whatever disease one is modelling. A benefit of the XGBoost model approach is that very good predictive properties are made possible automatically. The generalizability is difficult to assess, studies across other datasets, preferably from multiple countries would be valuable.

In conclusion the XGBoost machine learning model shows very promising results in evaluating risk of MI in a large and diverse population. This model can be used both for individual assessments and in larger cohorts.

## Data Availability

All data produced in the present study are available upon reasonable request to the authors

## Acknowledgments

We wish to thank the UK Biobank for making this research possible.

## Ethical approval

The North West Multi-Centre Research Ethics Committee approved the UK Biobank study. All participants provided written informed consent to participate. This research has been conducted using the UK Biobank Resource under Application Number 54045.

## Conflict of Interests

None

## Funding

None

## Contributorship statement

## Author Contributions

Mr Moore had full access to all the data in the study and takes responsibility for the integrity of the data and the accuracy of the data analysis.

*Concept and design:* Moore and Bell.

*Acquisition, analysis, or interpretation of data:* Moore and Bell.

*Drafting of the manuscript:* Moore and Bell.

*Critical revision of the manuscript for important intellectual content:* Moore and Bell.

*Statistical analysis:* Moore

*Obtained funding:* N/A.

*Supervision:* Bell.

## Supplementary Materials

### Logistic Regression

The goal of supervised regression models is to predict a target variable from a D-dimensional input vector. Linear regression models (such as the model implemented in this paper) will only use linear combinations of the input variables. In the case of two-class classification the target variable is written as a logistic sigmoid acting on a linear function of the input vector, maximum likelihood estimation is then used to determine the parameters of the logistic regression model^23^.

This paper used Scikit-learn’s implementation of a logistic regression model, with L2 regularization and ‘lbfgs’ solvers.

## Notes

### Competing Interest Statement

The authors have declared no competing interest.

### Funding Statement

This study did not receive any funding

